# Changes in Medicaid GLP-1 Prescription Dispensing After 4 States Ended Weight-Management Coverage

**DOI:** 10.64898/2026.11.15.26368150

**Authors:** Wei-Lun Lo, Rena M. Conti, Thuy Nguyen, Sumedha Gupta, Coady Wing, Kosali Simon

**Author notes:** Corresponding author: Kosali Simon, PhD.

## Abstract

On January 1, 2026, California, New Hampshire, Pennsylvania, and South Carolina ended Medicaid coverage of GLP-1s indicated for weight management. In 2025 Q4 these four states accounted for 60.5% of Medicaid weight-management GLP-1 prescriptions nationally (661,478 of 1,094,191). We used State Drug Utilization Data from 2024 Q1 through 2026 Q1 to describe dispensing in the first quarter after withdrawal, classifying products by FDA-approved indication. In the withdrawing states, weight-management GLP-1 prescriptions were 1.06 per 100 enrollee-months in 2025 Q3, 1.08 in 2025 Q4, and 0.16 in 2026 Q1, a decline of 85%, while in six states that kept coverage the same figures were 0.90, 0.97, and 0.82. The difference-in-differences estimate was −0.67 (95% CI, −1.21 to −0.18). Gross reimbursement in the withdrawing states fell by $1,079 per 100 enrollee-months from 2025 Q3 to 2026 Q1. Dispensing of GLP-1s labeled for other indications did not rise: the estimate against 33 never-covering states was −0.03 (95% CI, −0.29 to 0.23), so increases larger than 0.23 per 100 enrollee-months, about one quarter of the weight-management decline, can be ruled out. Substitution toward diabetes-labeled products was at most limited in the first quarter after coverage ended.

## Introduction

Glucagon-like peptide-1 receptor agonists (GLP-1s) are approved for weight management, diabetes, and other conditions. Federal law lets state Medicaid programs exclude drugs used for weight loss but requires coverage for other medically accepted uses.^1^

Some GLP-1s have weight-management-only U.S. Food and Drug Administration (FDA)-approved indications. Since December 2021, 17 states have opted to cover weight-management-indicated GLP-1s in their Medicaid program and coverage substantially increases utilization and spending.^2,3^ Citing budget pressure, California, New Hampshire, Pennsylvania, and South Carolina rescinded coverage of weight-management-indicated GLP-1s on January 1, 2026.^1,2^ In 2025 Q4, these four states accounted for 60.5% of Medicaid weight-management GLP-1 prescriptions nationally (661,478 of 1,094,191). We describe state Medicaid GLP-1 dispensing in the first quarter after coverage withdrawal, by FDA-approved indication.

## Methods

We used State Drug Utilization Data (SDUD) from 2024 Q1 through 2026 Q1, which report quarterly dispensed prescriptions and gross reimbursement by state, national drug code, and fee-for-service or managed care.^4^ We classified products by FDA-approved indication following prior work: weight-management (Saxenda, Wegovy, Zepbound, and generic liraglutide for weight management) or other indications (eMethods), and we set suppressed cells (fewer than 11 prescriptions per state, quarter, and product) to 5.^2^ Outcomes were prescriptions per 100 enrollee-months and gross reimbursement, which excludes rebates; enrollee-months are from CMS enrollment data.^5^

For weight-management GLP-1s, the comparison group was six states (Delaware, Kansas, Minnesota, Mississippi, Virginia, Wisconsin) that covered these products continuously from 2024 Q1 to 2026 Q1; states that changed coverage in the window and detailed handling of all states are in eMethods and a public repository.^6^ For non-weight-management GLP-1s, the comparison group was the 33 states that never covered weight-management GLP-1s; alternatively, we used the six kept-coverage states.^2^ We estimated difference-in-differences models with state and quarter fixed effects for 2024 Q1 to 2026 Q1, unweighted so that each state counts equally, as in prior work.^7^ Enrollment-weighted estimates are a sensitivity analysis. Inference used permutation, reassigning withdrawal across states. As a seasonal placebo, we repeated the analysis for 2024 Q3 to 2025 Q1, excluding South Carolina (which began coverage in November 2024) from the weight-management placebo (Table 1).

**Table 1.** Difference-in-differences estimates and sensitivity analyses.

|  | Estimate | 95% CI | P |
|---|---|---|---|
| **Weight-management GLP-1s (vs 6 kept-coverage states)** |  |  |  |
| Main estimate | −0.67 | −1.21 to −0.18 | 0.01 |
| *Alternative specification* |  |  |  |
| Two-period, 2025 Q3 to 2026 Q1 | −0.81 | −1.38 to −0.18 | 0.02 |
| Enrollment-weighted | −0.68 | −1.80 to 0.37 | 0.13 |
| *Alternative sample* |  |  |  |
| Excluding South Carolina (CA, NH, PA) | −0.81 | −1.37 to −0.33 | 0.01 |
| Adding Rhode Island to comparison states | −0.67 | −1.15 to −0.24 | 0.01 |
| *Placebo* |  |  |  |
| Seasonal placebo, 2024 Q3-2025 Q1 (CA, NH, PA) | 0.09 | −0.16 to 0.39 | 0.45 |
| **Non-weight-management GLP-1s (vs 33 never-covering states)** |  |  |  |
| Main estimate | −0.03 | −0.29 to 0.23 | 0.82 |
| *Alternative specification* |  |  |  |
| Enrollment-weighted | 0.10 | −0.37 to 0.48 | 0.56 |
| *Alternative sample* |  |  |  |
| 6 kept-coverage states as comparison | −0.04 | −0.32 to 0.21 | 0.80 |
| *Placebo* |  |  |  |
| Seasonal placebo, 2024 Q3-2025 Q1 (all 4 states) | −0.04 | −0.20 to 0.12 | 0.49 |

Estimates are prescriptions per 100 enrollee-months, unweighted unless noted, from models with state and quarter fixed effects. Except for the two-period and seasonal placebo rows, each is the 2026 Q1 change relative to each state's 2024 Q1-2025 Q3 mean, with 2025 Q4 entered separately. The two-period row compares 2026 Q1 with 2025 Q3 only. The seasonal placebo rows measure the change from 2024 Q3 to 2025 Q1, with 2024 Q4 entered separately; the weight-management placebo excludes South Carolina, which began coverage in November 2024. The kept-coverage row for non-weight-management-indicated GLP-1s uses the 6 kept-coverage states in place of the 33 never-covering states. CIs and P values are from permutation inference reassigning withdrawal across states (test inversion for CIs). Rows with 3 withdrawing states have 84 possible assignments, so the smallest attainable P is about 0.01.

This study used public, aggregate data and did not constitute human participants research.

## Results

In withdrawing states, weight-management GLP-1 prescriptions were 1.06 per 100 enrollee-months in 2025 Q3, 1.08 in 2025 Q4, and 0.16 in 2026 Q1, a decline of 85% from 2025 Q3, compared with 0.90, 0.97, and 0.82 in comparison states (Figure 1). The difference-in-differences estimate was ™0.67 (95% CI, ™1.21 to ™0.18). California accounted for 77% of the decline and Pennsylvania for 22%; South Carolina had little use before withdrawal (Figure 1). Gross reimbursement in withdrawing states fell by $1,079 per 100 enrollee-months from 2025 Q3 to 2026 Q1. The difference-in-differences estimate is smaller than the observed fall because it is measured against each state’s 2024 Q1 to 2025 Q3 average, when use was still rising. The seasonal placebo showed no significant differential change (estimate, 0.09; 95% CI, ™0.16 to 0.39).

**Figure 1.**
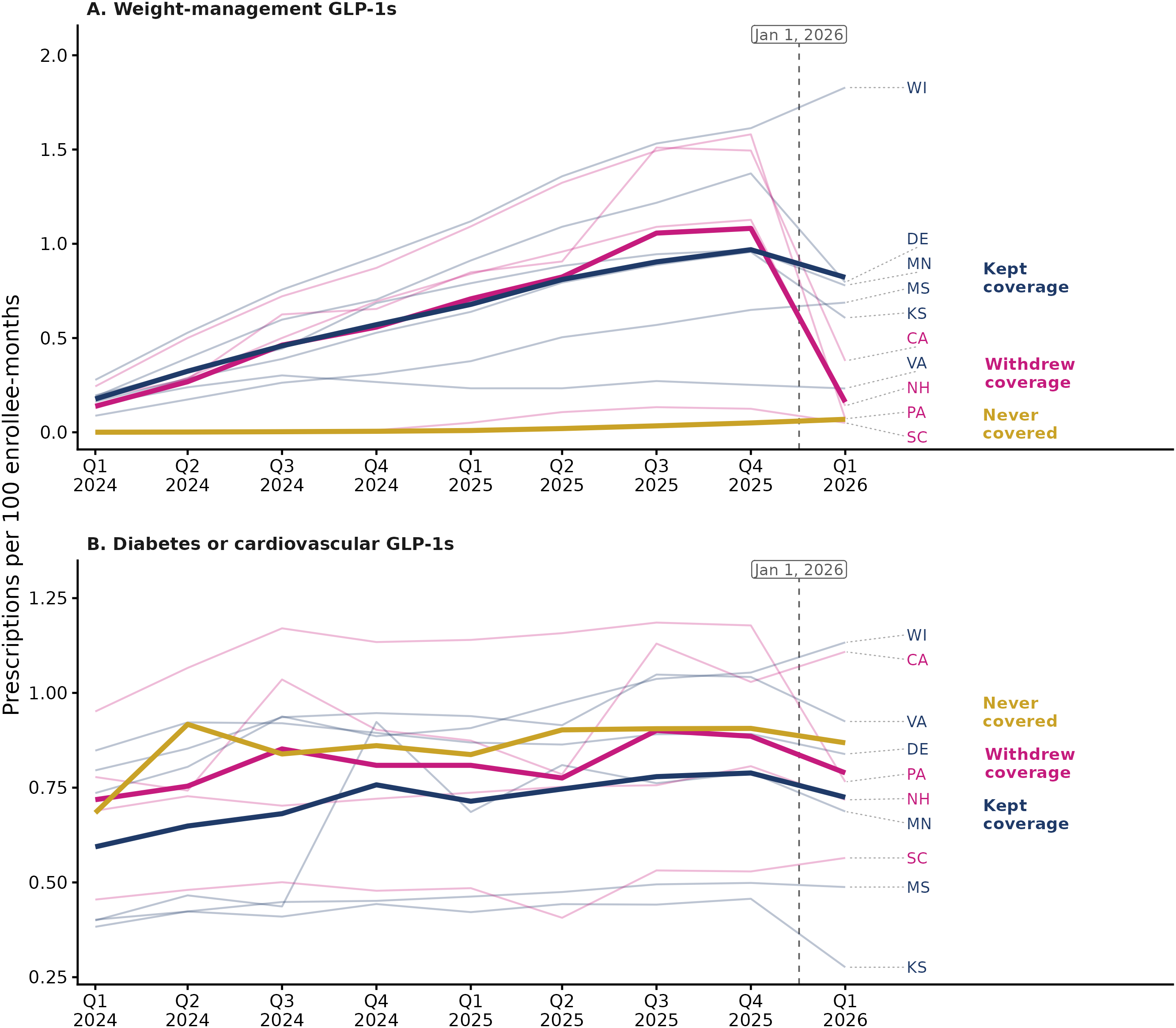
Medicaid GLP-1 prescriptions by 2026 coverage decision. Prescriptions per 100 Medicaid enrollee-months by quarter, 2024 Q1 to 2026 Q1, from State Drug Utilization Data. Thick lines show unweighted means across states in each group; thin lines show individual states (labeled at right); quarterly values are plotted at mid-quarter. Withdrawing states (4: California, New Hampshire, Pennsylvania, South Carolina) ended coverage of weight-management GLP-1s on January 1, 2026 (dashed line). Kept-coverage states (6: Delaware, Kansas, Minnesota, Mississippi, Virginia, Wisconsin) covered weight-management GLP-1s without interruption during the period; never-covering states (33) never covered them. Panel A: weight-management GLP-1s; the analysis compares withdrawing with kept-coverage states. Panel B: GLP-1s labeled for diabetes or cardiovascular disease and not for weight management (Wegovy, also labeled for cardiovascular risk reduction, is in Panel A); the analysis compares withdrawing with never-covering states.

In withdrawing states, non-weight-management prescriptions changed from 0.90 to 0.79 per 100 enrollee-months, compared with 0.91 to 0.87 in never-covering states (Figure 1; estimate, ™0.03; 95% CI, ™0.29 to 0.23), so we can rule out increases larger than 0.23 per 100 enrollee-months, about one quarter of the weight-management decline of 0.9. The estimate was similar with kept-coverage states as the comparison (™0.04; 95% CI, ™0.32 to 0.21); the largest rise in any withdrawing state was 0.03, and the seasonal placebo estimate was ™0.04 (95% CI, ™0.20 to 0.12).

## Discussion

In the first quarter after four states ended Medicaid coverage of weight-management GLP-1s, dispensing of these products fell by about 85%, while dispensing in states that kept coverage changed little. Dispensing did not fall by 100%, consistent with continued coverage for enrollees under 21 and residual claims from before the change.^8,9^

Prescribing of non-weight-management-indicated GLP-1s did not rise in withdrawing states; any increase consistent with the data is small relative to the weight-management decline, so substitution was at most limited. Prior studies of coverage adoption reached mixed conclusions on whether weight-management and other GLP-1 use substitute for one another;^2,7^ withdrawal offers a complementary test.

The study has several limitations: SDUD contain no diagnoses, so products are classified by labeled indication; off-label use of diabetes-labeled products for weight loss is not observed, but California, Pennsylvania, and South Carolina Medicaid required a type 2 diabetes diagnosis for those products.^10^ We only had access to a single post-withdrawal quarter of data, which may be revised as states submit late claims.^4^ Comparison states adopted coverage at different times and may still have been on their own post-adoption paths.

Person-level claims data are needed to assess whether people who lost weight-management-indicated GLP-1 coverage stopped treatment, paid out of pocket, or switched products, and the consequences for their health and spending.

## Supporting information

ESupplement

## Data Availability

State Drug Utilization Data and CMS Medicaid enrollment data are public. The state coverage evidence file, change log, criteria, and the list of 251 GLP-1 NDCs are in a public GitHub repository (https://github.com/simonkos-cell/state-medicaid-glp1-coverage-rules). The replication code is in a separate repository that will be made public on acceptance.

