## Supplementary material for "Changes in Medicaid GLP-1 Prescription Dispensing After 4 States Ended Weight-Management Coverage": ESupplement

This supplemental material has been provided by the authors to give readers additional information about their work.

**eMethods.** GLP-1 product classification, measures, and state coverage groups

**eReferences**

### eMethods. GLP-1 Product Classification, Measures, and State Coverage Groups

#### Product classification

State Drug Utilization Data (SDUD) contain no diagnoses, so each national drug code (NDC) was classified by the US Food and Drug Administration–approved indications of the product, following prior work.[1] The list of 251 GLP-1 NDCs is in the replication repository.[2]

*Weight-management:* Wegovy (semaglutide), Saxenda (liraglutide), generic liraglutide labeled for weight management, and Zepbound (tirzepatide).

*Other indications:* Ozempic and Rybelsus (semaglutide); Victoza and generic liraglutide labeled for diabetes; Trulicity (dulaglutide); Mounjaro (tirzepatide); Byetta, Bydureon, and generic exenatide; Soliqua (lixisenatide and insulin glargine); and Xultophy (liraglutide and insulin degludec). Adlyxin, Tanzeum, and 1 unbranded semaglutide NDC are on the list but have no SDUD records in 2024 Q1 to 2026 Q1.

Wegovy is also labeled for cardiovascular risk reduction and metabolic dysfunction–associated steatohepatitis, and Zepbound for obstructive sleep apnea. Both remain in the weight-management group, so that group includes any prescriptions for these indications, which the 2026 withdrawals did not end.

#### Measures

The withdrawing states' share of Medicaid weight-management prescriptions, reported in the Introduction, is their 2025 Q4 weight-management prescription count divided by the count for all 50 states.

#### State coverage groups

Coverage dates come from state preferred drug lists, prior-authorization criteria, state plan amendments, and Medicaid bulletins, assembled for Wing et al[1] and audited against the source documents in October 2026. The evidence file, change log, and criteria for each state are public.[3] The table that follows assigns each of the 17 covering states to a group.

A state qualified as a kept-coverage comparison if it covered at least 1 weight-management GLP-1 in every quarter from 2024 Q1 through 2026 Q1 and neither started nor ended coverage in that window. Changes short of starting or ending coverage, listed in the table, did not disqualify a state. Virginia (2024 Q3, BMI threshold raised), Wisconsin (July 2024, criteria revised), and Kansas (April 2024 and June 2025, BMI-40 tier waived for Zepbound and Wegovy) adjusted prior-authorization criteria while covering continuously; all preceded 2025 Q3, the base quarter of the two-period estimate, but fell within the main model's baseline, and the adjustments ran in opposite directions. States whose coverage started, ended, or was interrupted in the window (MA, MO, NC, TN, UT) or was restricted in January 2026 (MI) were excluded.

Kansas is the one comparison state whose documented start falls inside the window. Its April 2024 date marks Zepbound becoming preferred; SDUD show Saxenda paid from 2019 and Wegovy by 2021 Q4, with both paid in 2024 Q1, so Kansas was treated as covering throughout. Rhode Island covered throughout the window but ended coverage after the last quarter analyzed, so it enters only as a sensitivity analysis.

The other-indication comparison uses the 33 states that never covered a weight-management GLP-1: AK, AL, AR, AZ, CO, CT, FL, GA, HI, IA, ID, IL, IN, KY, LA, MD, ME, MT, ND, NE, NJ, NM, NV, NY, OH, OK, OR, SD, TX, VT, WA, WV, and WY. These include Alabama, Hawaii, Louisiana, and Washington, which cover Wegovy only for cardiovascular risk reduction, and West Virginia, which covers Zepbound only for obstructive sleep apnea.

#### Coverage of Weight-Management GLP-1s in the 17 Covering States

| State | Coverage start | Changes, 2024 Q1 to 2026 Q1 |
|---|---|---|
| **Withdrawing states** | | |
| California | January 2023 | Zepbound added October 2024; ended January 1, 2026 |
| New Hampshire | September 2022 | Zepbound added September 2024; ended January 1, 2026 |
| Pennsylvania | January 2023 | Zepbound added January 2024; ended January 1, 2026 |
| South Carolina | November 2024 | Began in window (Zepbound not covered); ended January 1, 2026 |
| **Kept-coverage comparison states** | | |
| Delaware | January 2023 | Zepbound added August 2024; state plan amendment on comorbidity and prior authorization effective August 2024 |
| Kansas | April 2024 (documented); Saxenda paid from 2019 | Zepbound preferred with BMI 40 tier waived, and prior authorization required for Saxenda and Wegovy, April 2024; BMI 40 tier waived for Wegovy, June 2025 |
| Minnesota | December 2021 | Zepbound added by January 2025 |
| Mississippi | July 2023 | None (Zepbound not covered) |
| Virginia | January 2023 | BMI threshold raised, 2024 Q3; August 2024 list adds Zepbound, with Saxenda preferred and Wegovy and Zepbound nonpreferred |
| Wisconsin | April 2023 | Zepbound added and clinical criteria revised, July 2024 |
| **Sensitivity analysis only** | | |
| Rhode Island | October 2022 | Zepbound added May 2024; ended October 1, 2026, after the window |
| **Excluded** | | |
| Massachusetts | January 2024 | Adult Wegovy and Saxenda ended January 1, 2025; new criteria February 17, 2026 |
| Michigan | February 2022 | Zepbound added November 2024; restricted to BMI 40 or higher, January 1, 2026 |
| North Carolina | August 2024 | Began in window; ended October 1, 2025; reinstated December 12, 2025 |
| Missouri | October 2024 to February 2025 | Began in window |
| Tennessee | August 2025 | Began in window |
| Utah | July 2025 | Began in window |

Abbreviation: BMI, body mass index (calculated as weight in kilograms divided by height in meters squared).

Coverage start is the earliest date for any of Wegovy, Saxenda, or Zepbound in the evidence file,[3] generally the first document in hand listing the class. SDUD show Saxenda paid earlier in California, Delaware, Kansas, New Hampshire, Rhode Island, Virginia, and Wisconsin, so these dates are lower bounds; only the Kansas date falls inside the window.
